# The Finger Dexterity Test: validation study of a smartphone-based manual dexterity assessment

**DOI:** 10.1101/2023.06.20.23291590

**Authors:** Delphine Van Laethem, Stijn Denissen, Lars Costers, Annabel Descamps, Johan Baijot, Ann Van Remoortel, Annick Van Merhaegen-Wieleman, Marie B D’hooghe, Miguel D’Haeseleer, Dirk Smeets, Diana M Sima, Jeroen Van Schependom, Guy Nagels

**Author notes:** **Corresponding author:** Name: Delphine Van Laethem, Email address. Shared first authors (Delphine Van Laethem, Stijn Denissen).

## Abstract

**Background:** The Nine-Hole Peg Test (9HPT) is the golden standard in clinical practice to measure manual dexterity in people with multiple sclerosis (MS). However, administration requires trained personnel and dedicated time during a clinical visit.

**Objective:** To validate a smartphone-based test for remote manual dexterity assessment, the icompanion Finger Dexterity Test (FDT), to be included into the icompanion application.

**Methods:** 65 MS and 81 healthy subjects performed a first testing session, and 21 healthy subjects performed a second session approximately two weeks later.

**Results:** The FDT significantly correlated with the 9HPT (dominant: Spearman’s ρ=0.62, p<0.001; non-dominant: ρ=0.52, p<0.001). FDT scores showed a significant difference between the MS and healthy subjects (dominant: Cohen’s d=0.24, p=0.015; non-dominant: Cohen’s d=0.18, p=0.013), which was not the case for the 9HPT. A significant correlation with age (dominant: ρ=0.46, p<.001; non-dominant: ρ=0.40, p=0.002), Expanded Disability Status Scale (EDSS, dominant: ρ=0.37, p=0.004; non-dominant: ρ=0.33, p=0.017), and disease duration for the non-dominant hand (ρ=0.31, p=0.016) was observed.

**Conclusion:** The icompanion Finger Dexterity Test shows a moderate-to-good concurrent validity, ecological validity and test-retest reliability, and differentiates between the MS subjects and healthy controls. This test can be implemented into routine MS care for remote follow-up of manual dexterity.

## Introduction

Multiple sclerosis (MS) is the most common inflammatory and neurodegenerative disease in young adults, affecting 2.8 million people worldwide (1). Between 60 and 75% of persons with MS (PwMS) with mild disability have an impaired manual dexterity, with an increasing prevalence with worsening disease (2). This impairment is related to decreased strength, sensory dysfunction and/or tremor and results in a decreased participation in social, home and productive activities (3).

The Nine-Hole Peg Test (9HPT) is considered the golden standard for the assessment of manual dexterity in MS, both in a clinical and research context (4). In this test, subjects have to place nine pegs into nine holes and subsequently remove them one by one. This test has a good convergent, discriminant and ecological validity and test-retest reliability. However, its administration requires clinic visits, appropriate material and trained personnel, which not all clinical sites have the resources for. Furthermore, there are significant practice effects after repeated administration, resulting in a biased estimation of finger dexterity. Finally, MS is characterized by important fluctuations of symptoms due to temperature changes (5) and fatigue (6). The moment of a clinical visit, which in itself may cause stress and fatigue, might thus not be the most accurate representation of an individual’s function.

Since the COVID-19 pandemic, there has been an increasing interest in remote patient monitoring, which reduces the need for clinical visits, and thus costs and patient burden. Smartphone-based assessments are of particular interest due to their broad accessibility (7) and allow the patient to perform the evaluation at a time of their choosing.

In this study, we developed a smartphone-based test for the assessment of manual dexterity in multiple sclerosis and assessed the concurrent, ecological validity and test-retest reliability of the Finger Dexterity Test (FDT), and its ability to differentiate between PwMS and healthy controls.

## Methods

### Study protocol and ethics

The study protocol, patient information and informed consent were approved by the ethics committees at the University Hospital Brussels (B.U.N. 143201940335, 2019/173, 17 July 2019) and National MS Center Melsbroek. Informed consent was obtained from all participants before inclusion. The FDT validation is part of a bigger validation study of a smartphone app that assesses both cognitive performance and manual dexterity in PwMS, to be included into the icompanion app (8).

### Recruitment

PwMS were recruited at the University Hospital Brussels and the National MS Center Melsbroek, whereas healthy control subjects (HC) were recruited via online advertising, leaflets and acquaintances of the examiners-. All subjects were native Dutch-speaking or bilingual. For PwMS, the only additional inclusion criterion was a confirmed diagnosis of MS based on the modified McDonald criteria (9), whereas a subject was considered a healthy control if no neurological, psychiatric or learning disorder was present. Exclusion criteria for PwMS were the presence of other neurological,psychiatric or learning disorders, hospitalization in the context of a medical problem and a relapse within the last month.

In total, 65 PwMS and 81 HC were included in this study. Of those, 60 PwMS and 79 HC completed the FDT for both the dominant and non-dominant hand. To assess test-retest reliability, 21 HC were tested a second time 2-3 weeks later, on a similar moment to the first testing session (morning versus evening, weekday versus weekend). During this second session, only the smartphone-based tests were repeated.

### Demographic and clinical information

The following demographic information was collected: age, sex, education level, subjective visual and manual problems (yes/no), current medication, recent cognitive assessment and Edinburgh Handedness Inventory (EHI, (10)). In addition, the following information was collected for PwMS: EDSS score (assessed by a neurologist during clinical visit), disease duration and type.

### Finger Dexterity Test

All tests were performed on a preconfigured smartphone (Samsung Galaxy A10), on which an application was installed. In the Finger Dexterity Test, the goal is to “fry” and subsequently “serve” a total of 10 eggs as quickly as possible. For each of the eggs (presented on the same horizontal position in the screen but with a random permutation of 10 different vertical positions), a sequence of steps is performed. First, an egg is presented on the screen, which the subject has to break by pinching it between two fingers. In a second step, the subject needs to drag and release the egg to a frying pan, where it is cooked. Finally, the cooked egg is pinched and dragged to a plate, where it is served (see Figure 1). A practice phase of 10 eggs precedes the actual testing phase. The entire test, including both the practice and test phase, is performed first with the dominant hand and subsequently with the non-dominant hand. The final score is defined as the total time to “serve” 10 eggs, which is recorded by the application.

**Figure 1:**
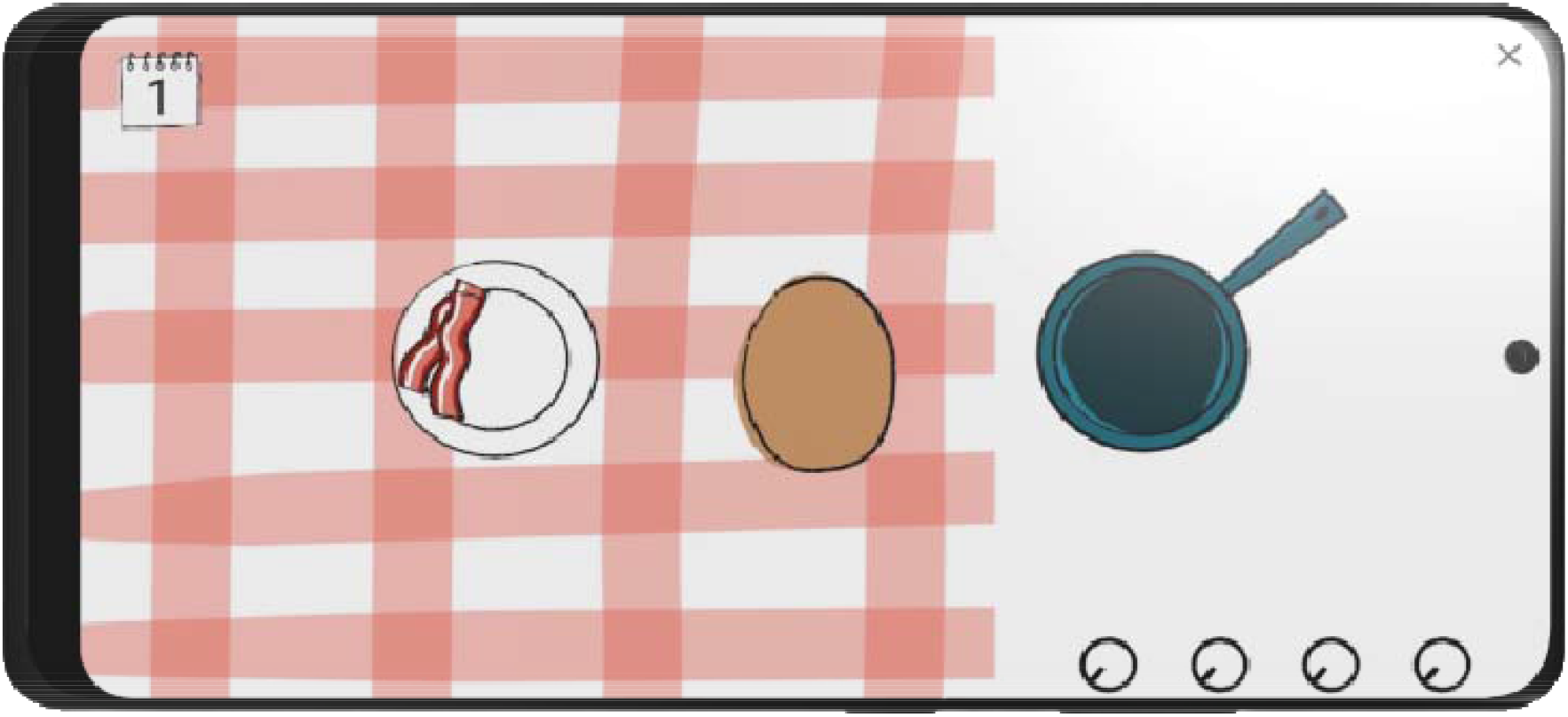
Finger Dexterity Test

### Nine-Hole Peg Test

The objective is to transfer nine pegs from a container into nine holes (phase 1) and subsequently back to the container (phase 2), one peg at a time, as quickly as possible. Subjects performed two trials with the dominant hand, followed by two trials with the non-dominant hand. If a peg fell on the table but was still reachable by the subject, the trial was continued. If the peg fell on the floor, the trial was discontinued and repeated from the start. The time to complete each trial was measured using a stopwatch. The score is the average time to complete the two trials for each hand (4).

### Questionnaires

At the end of the testing session, subjects were asked to complete the Beck Depression Inventory (BDI, (11)) and the Fatigue Scale for Motor and Cognitive functions (FSMC, (12)). Finally, the perceived burden of the performed tests was assessed on a 7-point scale. These questionnaires were completed independently by the subjects on paper, but they were able to ask questions or clarification if necessary.

### Statistics

Group differences between PwMS and HC were assessed with a Chi-square test for categorical variables and a Mann-Whitney-U test for continuous variables.

Concurrent validity was assessed using Spearman correlation between the FDT and the Nine-Hole Peg Test. The difference in mean FDT score between PwMS and HC were investigated using a Mann-Whitney-U test. Ecological validity was investigated by assessing the Spearman correlation between the FDT on the one hand and age, disease duration, EDSS, BDI score and FSMC score on the other. Finally, test-retest reliability was investigated in the subset of HC who performed a second testing session, by assessing the Spearman correlation between the FDT score at the first and second sessions.

To interpret the magnitude of concurrent and ecological validity, we used the classification of Portney et al. 2009 (13), with a correlation coefficient < 0.25, 0.25-0.50, 0.50-0.75 and > 0.75 being considered as small, fair, moderate to good and good to excellent respectively. The interpretation of the magnitude of test-retest reliability was based on the classification Koo et al. 2016 (14), with a coefficient < 0.5, 0.5-0.75, 0.75-0.9, and > 0.90 indicating a poor, moderate, good, and excellent reliability respectively.

Per validation criterion, we performed an additional analysis without FDT outliers. Outliers were only excluded from the analyses if they met both of the following conditions: value below the 25^th^ percentile minus 1.5 times the interquartile range or above the 75^th^ percentile plus 1.5 times the interquartile range, and a valid reason for exclusion (i.e. if there was a lag of the application or interference of long fingernails).

Finally, to correct performance on the FDT for age and sex, we performed a regression-based normalisation separately for the dominant and the non-dominant hand, which can be found in Supplementary Materials.

A two-sided test with a type I error probability of 0.05 was used for all analyses.

## Results

### Demographic

In table 1, the group differences between PwMS and HC are depicted. PwMS had a higher score on the questionnaires for depression (BDI) and fatigue (FSMC). There was no significant difference between PwMS and HC on any of the other demographic and clinical parameters.

**Table 1:**
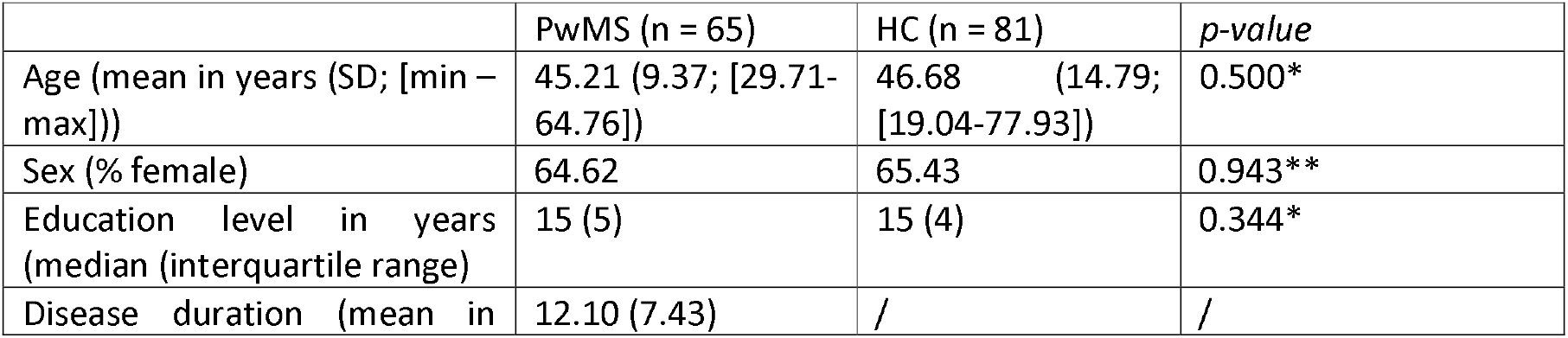
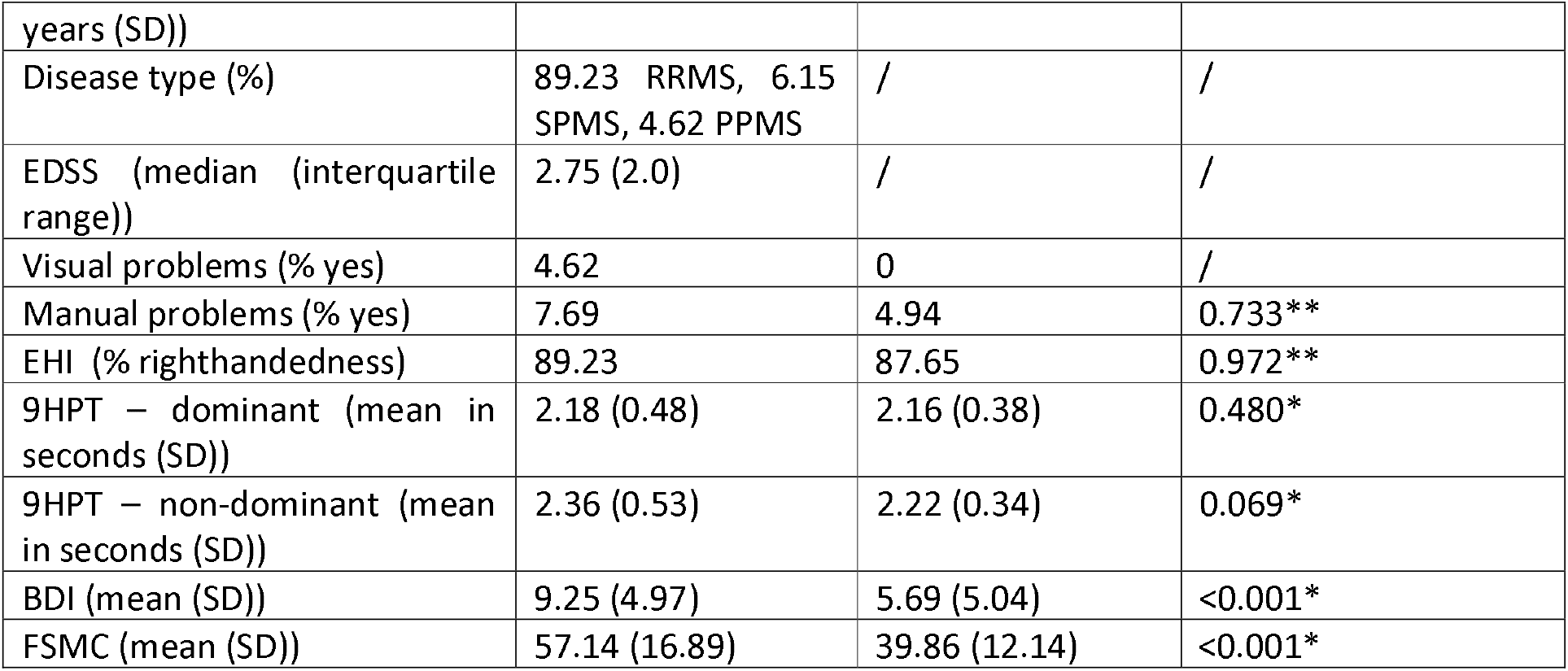
Demographic and clinical information-RRMS = relapsing-remitting MS, SPMS = secondary progressive MS, PPMS = primary progressive MS, SD = standard deviation, min = minimum value, max = maximum value *Mann-Whitney U test, ** Chi-Squared test

### Concurrent validity

There was a significant correlation between the 9HPT and FDT scores of the PwMS in the dominant and non-dominant hand (Spearman’s ρ (hereafter referred to as ρ) = 0.62, p < 0.001 and ρ = 0.52, p < 0.001 respectively), as well as the HC (dominant hand: ρ = 0.28, p = 0.012; non-dominant hand: ρ = 0.37, p = 0.001). When excluding outliers (2 dominant, 1 non-dominant), the correlations with the 9HPT remained significant for the PwMS (dominant: ρ = 0.63, p < 0.001; non-dominant: ρ = 0.49, p < 0.001) and the HC (dominant: ρ = 0.30, p = 0.007; non-dominant: ρ = 0.35, p = 0.002). The association between these two variables is depicted in the Figure 2 and 3. The depiction of without outliers can be found in Supplementary Materials (Figure S1 & S2).

**Figure 2.**
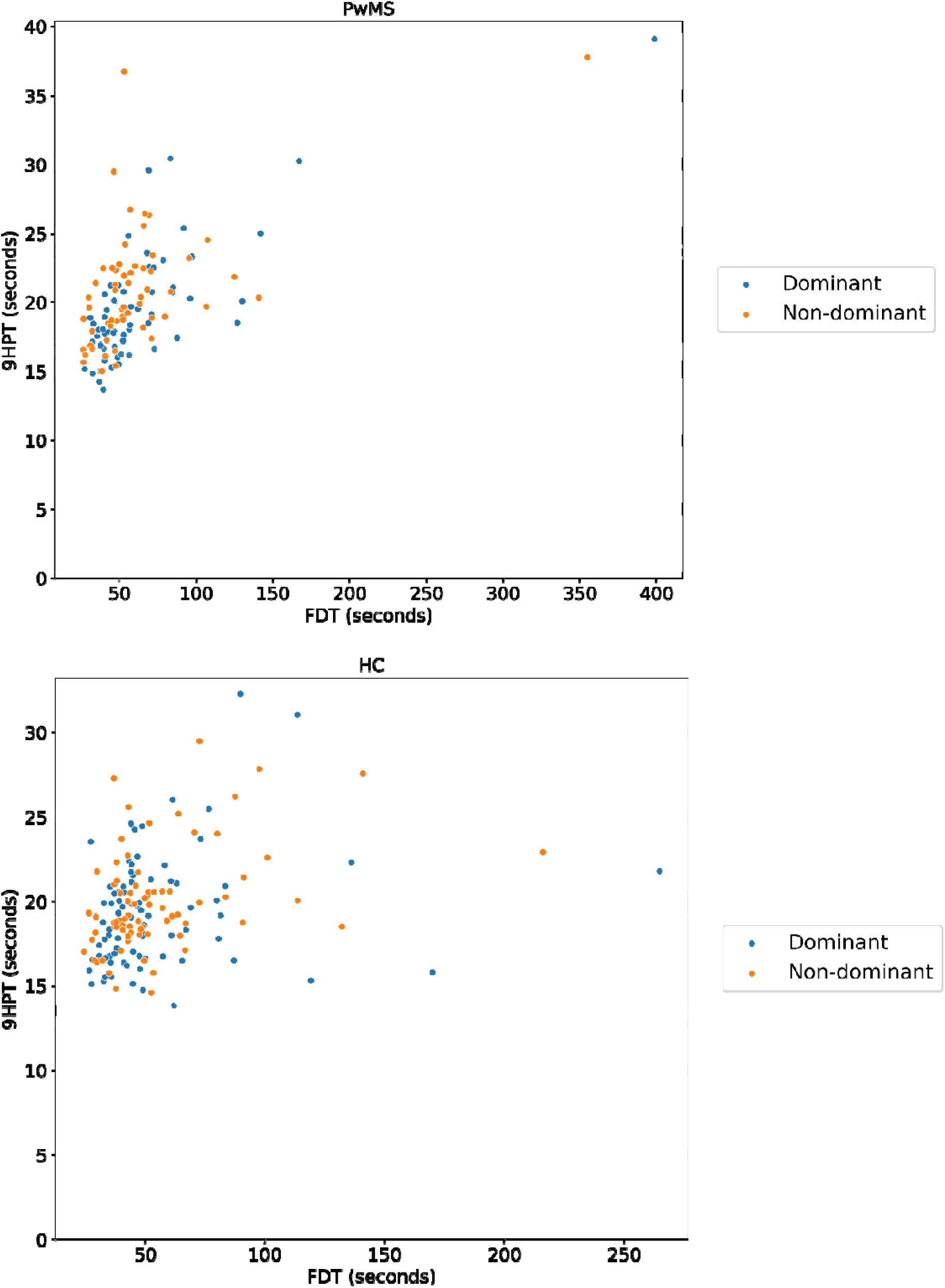
&3: Concurrent validity in subjects with MS (PwMS) and healthy control subjects (HC) -scatterplot between Finger Dexterity Test (FDT) and Nine-Hole Peg Test (9HPT).

### Capacity to differentiate between MS and HC

There was a significant difference in FDT between PwMS and HC, both in the dominant and non-dominant hand. The difference between the two groups is depicted in Figure 4 & 5 and Table 2. When removing FDT outliers (MS: 2 dominant hand, 1 non-dominant hand; HC: 2 dominant hand, 5 non-dominant hand), the group difference remained significant for both hands (dominant: Cohen’s d = 0.33, p = 0.014; non-dominant: Cohen’s d = 0.33, p = 0.004). The depiction without outliers can be found in Supplementary Materials (Figure S3 & S4).

**Table 2:** Group difference of Finger Dexterity Test (FDT) between PwMS and HC, reported as means with standard deviations. * Mann-Whitney U test

|  | PwMS | HC | Cohen’s d | p-value |
| --- | --- | --- | --- | --- |
| FDT – dominant hand (in seconds) | 64.51 (50.26) | 54.43 (34.12) | 0.24 | 0.015* |
| FDT – non-dominant hand (in seconds) | 61.64 (45.52) | 55.02 (29.16) | 0.18 | 0.013* |

**Figure 4.**
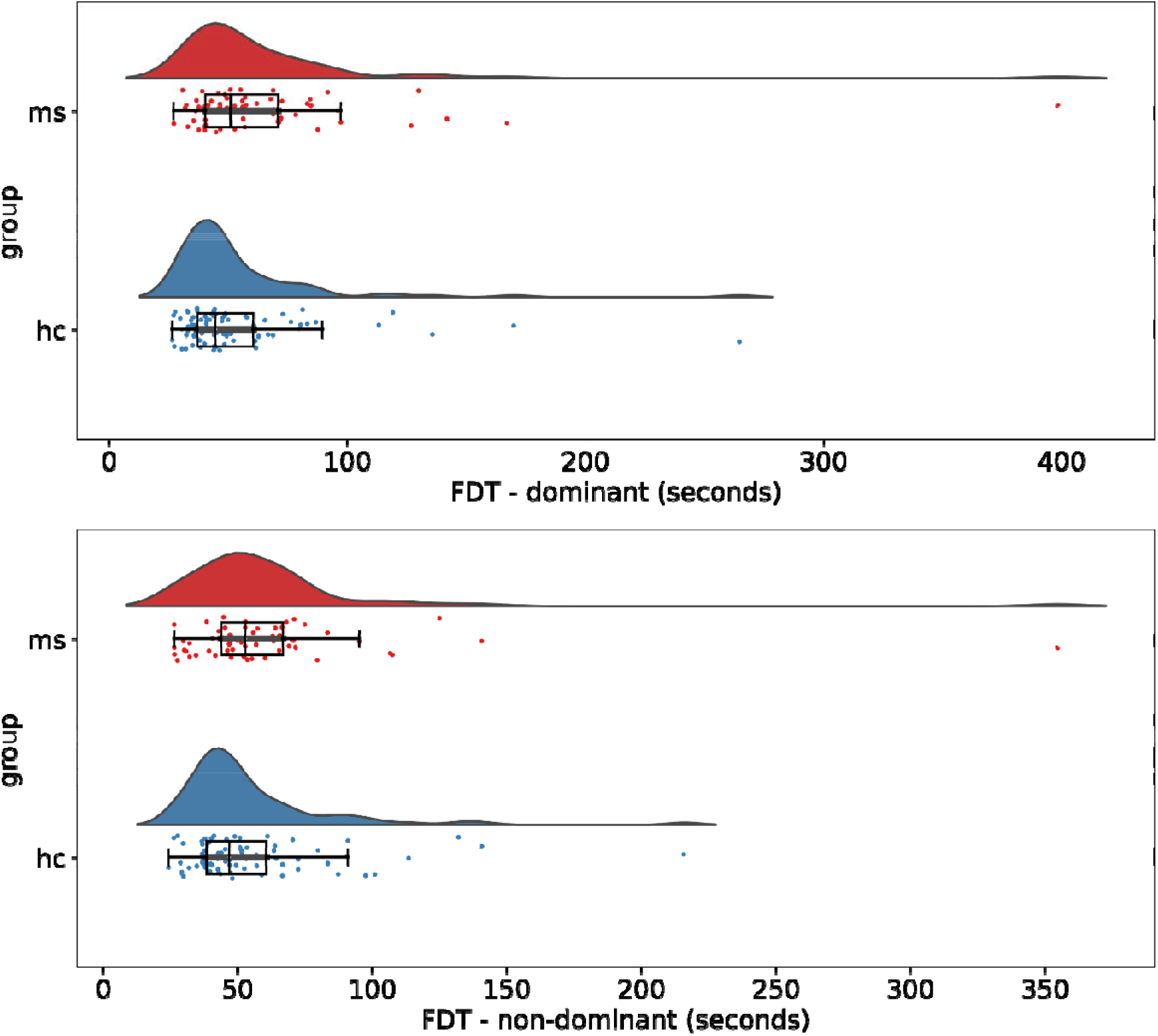
& 5: Group difference of Finger Dexterity Test (FDT) between subject with MS and healthy controls, dominant and non-dominant hand.

### Ecological validity

There was a significant correlation with age in the dominant and non-dominant hand in PwMS (ρ = 0.46, p < 0.001 and ρ = 0.40, p = 0.002 respectively) and HC (ρ = 0.42, p < 0.001 and ρ = 0.41, p < 0.001 respectively). The correlation with EDSS was also significant for both hands (ρ = 0.37, p = 0.004 and ρ = 0.33, p = 0.018). Finally, the correlation with disease duration was only significant for the non-dominant hand (ρ = 0.31, p = 0.016). When removing FDT outliers (2 dominant hand, 1 non-dominant hand), all correlations remained significant (age PwMS: dominant ρ = 0.50, p < 0.001 & non-dominant: ρ = 0.40, p = 0.002; age HC: dominant ρ = 0.39, p < 0.001 & non-dominant: ρ = 0.40, p < 0.001; EDSS: dominant ρ = 0.40, p = 0.002 & non-dominant ρ = 0.30, p = 0.036; disease duration: non-dominant ρ 0.29, p = 0.032). Other correlations with education and measures of depression (BDI) and fatigue (FSMC) were not significant in PwMS. In HC, there was a significant correlation with FSMC in the non-dominant hand (ρ = -0.29, p = 0.015), which remained significant after removing FDT outliers (ρ = -0.31, p = 0.009). The associations with age, EDSS and disease duration in the PwMS are depicted in Figure 6, 7 & 8.

**Figure 6, 7 & 8:**
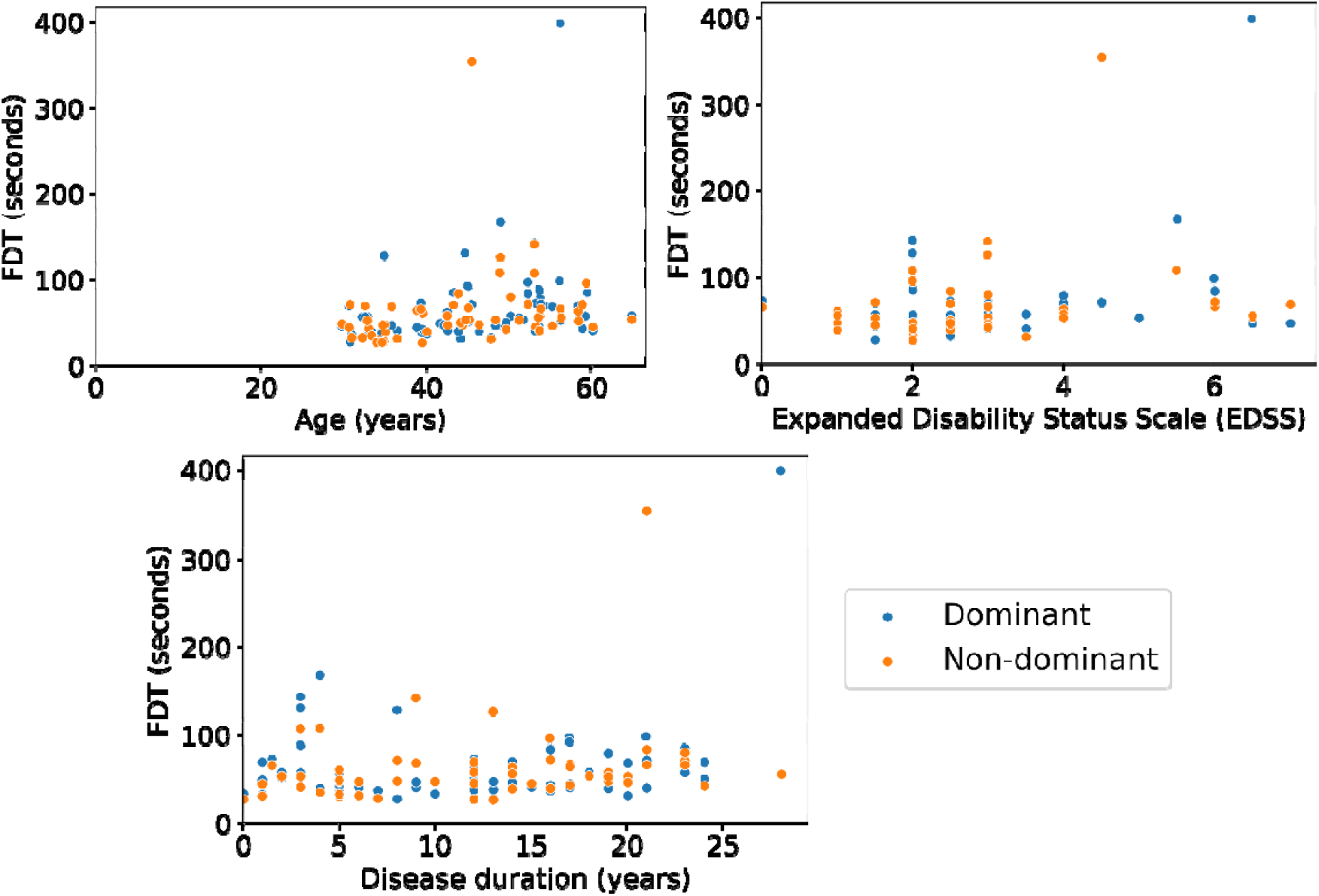
Ecological validity - scatterplot between Finger Dexterity Test (FDT) and age, EDSS and disease duration.

### Test-retest reliability

There was a significant association, both in the dominant and non-dominant hand (ρ = 0.71, p < 0.001 and ρ = 0.82, p < 0.001), between the FDT score at the first and second testing sessions for the HC. The association between the first and second testing session is depicted in Figure 9. When removing FDT outliers (0 for dominant hand, 5 for non-dominant hand), the correlations remained significant for both hands (ρ = 0.66, p = 0.002 and ρ = 0.91, p < 0.001).

**Figure 9:**
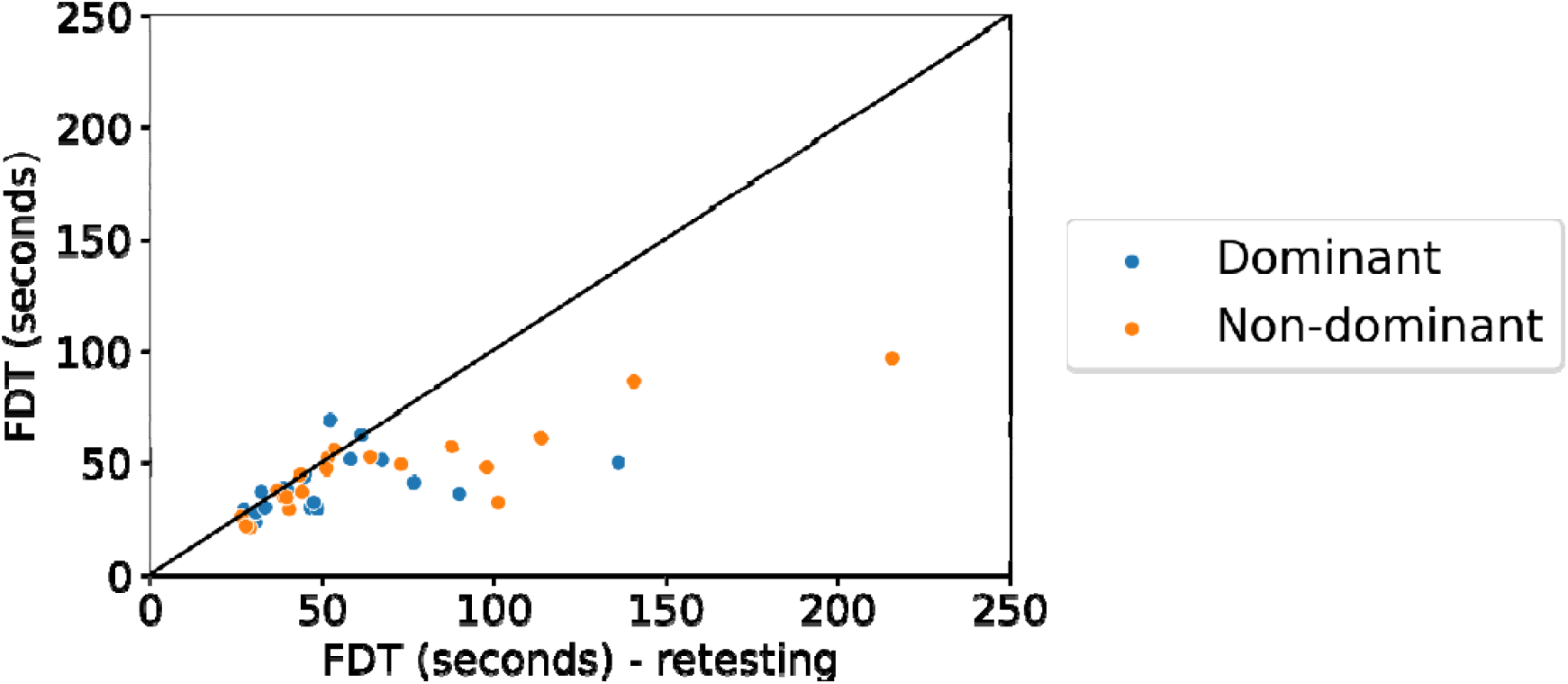
Test-retest reliability of the Finger Dexterity Test (FDT) in healthy controls: scatterplot between Finger Dexterity Test (FDT) scores of the first and second testing session, with identity line (x = y).

## Discussion

In this study, we established the validity of the smartphone-based icompanion Finger Dexterity Test. The test met all validation criteria: there was a moderate-to-good concurrent validity with the 9HPT (dominant hand: ρ = 0.62, p < 0.001; non-dominant hand: ρ = 0.52, p < 0.001) and an ability to distinguish between PwMS and HC (Cohen’s d = 0.24, p = 0.015 for the dominant hand and Cohen’s d = 0.18, p = 0.013 for the non-dominant hand). Furthermore, there was a weak-to-moderate ecological validity in MS subjects (age: ρ = 0.46, p < .001 for dominant hand and ρ = 0.40, p = 0.002 for non-dominant hand; EDSS: ρ = 0.37, p = 0.004 for dominant hand and ρ = 0.33, p = 0.017 for non-dominant hand; disease duration ρ = 0.31, p = 0.016 for non-dominant hand) and a good-to-excellent test-retest reliability in HC (ρ = 0.71, p < 0.001 for the dominant hand, ρ = 0.82, p < 0.001 for the non-dominant hand).

In a post-hoc analysis we used an alternative scoring method, namely the number of eggs/second instead of the total time of serving 10 eggs, as suggested in Feys et al. 2017 to reduce floor effects and allow for a more normal distribution of the data (4). The above-mentioned correlations remain similar with this method, but the group difference between PwMS and HC is no longer significant for the non-dominant hand (Cohen’s d = -0.36, p = 0.015 for the dominant hand and Cohen’s d = -0.17, p = 0.31 for the non-dominant hand).

Of note, there was no significant difference between PwMS and HC for the 9HPT, which is the golden standard for the assessment of manual dexterity in MS (4). This may be explained by the frequent assessments carried out in the tertiary centers where the MS subjects were recruited. On the other hand, the included subjects had a relatively mild degree of disability (median EDSS of 2.75, with only around 11% having a progressive disease course), and may thus not yet show impairment of manual dexterity on the 9HPT. In any case, this warrants caution when using manual dexterity assessments frequently in clinical practice.

The Floodlight application (15) contains similar tests for assessing upper extremity function in MS: In the Pinching Test, tomatoes appearing on the screen have to be squeezed between two fingers, thereby evaluating fine pinching and grasping dexterity. Scoring is based on the number of tomatoes squeezed within 30 seconds, and the gap between the first and second finger touching the screen. In the Draw a Shape Test subjects have to draw certain shapes on the screen to evaluate fine finger or manual dexterity. Scoring is based on accuracy and the ratio of accuracy and time needed to draw the shape (15). They found a moderate to good test-retest reliability and concurrent and ecological validity. Furthermore, both tests provided independent information in the prediction of the 9HPT (15). While both the Floodlight tests and the FDT show a similar concurrent validity and test-retest reliability (15), the FDT has the advantage that only one test needs to be performed by the patient to evaluate manual dexterity. By not subdividing the task into different subcomponents, our test may be more representative of the manual dexterity used in daily real-world activities, as multiple aspects of dexterity are used in daily life tasks. Furthermore, scoring is based on the total time needed to complete the test and is, therefore easier to interpret than the multiple subscores provided by the Floodlight tests. Finally, the FDT more closely resembles the 9HPT, as both are characterized by a similar pinch and drag/transfer movement. This might help with the clinical translatability of the FDT scores, and consequentially the adoption of the assessment in clinical practice.

The implementation of the FDT into daily practice poses many advantages. First, manual dexterity assessment can be carried out independently by the patient at home as a regular screening for (the progression of) manual dexterity problems. This reduces the need for testing during clinical visits. This reduces the duration of the visit and the need for trained personnel. Moreover, patients can take the frequent symptom fluctuations (5,6) better into account and perform the test at the most opportune time. Furthermore, the test is rater-independent, and allows for the patient to take a more active role in their own clinical follow-up. In the future it could be interesting the assess the relationship between FDT scores and measures of self-reported difficulties in upper extremity function in MS, such as the Arm Function in Multiple Sclerosis Questionnaire-Short Form (AMSQ-SF, (16)). Finally, mobile applications have the potential to allow assessment of multiple aspects of the test performance, such as speed or difficulty of different aspects of the test (for example pinch versus drag movement)and fatigability (i.e. reduced task performance after sustained effort (17)). The investigation of these specific parameters is an important next step in the research of digital manual dexterity assessment tools, which may replace paper-pencil tests in the near future.

There are also limitations inherent to smartphone-based assessments that the patient carries out without supervision. First, differences in the execution of the test (phone in hand versus on table, pinch between thumb and second digit versus second and third digit) cannot be verified, as well as influencing factors such as distraction and long fingernails (capacitive touchscreens do not register touch commands performed with a fingernail because it is nonconductive (18)). To mitigate this, patients are able to report any issues after completion of the test. Second, experience interacting with digital technology such as smartphone apps can have an impact on the test performance, especially in older individuals, but this is difficult to assess. Furthermore, as with all neuroperformance tests, particularly cognitive and manual dexterity tests (19–21), it is important to take practice effects into account. To reduce this risk, a limit could be set to the test frequency. A study where patients perform the test in the home setting may be indicated to better assess the impact of these limitations and take appropriate countermeasures. Another limitation is testing was carried out on the same model of smartphone for all subjects. It is unclear how variations in screen size and touchscreen technology could influence test performance. Finally, the application lagged for some subjects during the testing phase of the non-dominant hand, for which we were unable to identify the underlying cause. Excluding the subjects for whom the score was an outlier and for whom the application lagged, did however not significantly influence the results.

## Conclusion

In conclusion, the Finger Dexterity Test is a valid and reliable tool to assess manual dexterity in MS, and could overcome practical issues with the current golden standard of assessing manual dexterity in daily practice.

## Data Availability

The source data and the code used for the statistical analysis and the creation of the figures will be made available in the GitHub repository of our lab, in a sub-repository named smartphone_tests.

https://github.com/AIMS-VUB/smartphone_tests

## Acknowledgements

We would like to thank all subjects who were willing to participate in our study for their invested time and effort. Furthermore, we would like to thank Florine Wöhler and Eva Keytsman for their invaluable help in subject testing, as well as Steve De Backer for his help in the creation of the application.

## Data availability statement

The source data and the code used for the statistical analysis and the creation of the figures will be made available in the GitHub repository of our lab, in a sub-repository named ‘smartphone_tests’: https://github.com/AIMS-VUB/smartphone_tests

