## Supplementary materials for "The Finger Dexterity Test: validation study of a smartphone-based manual dexterity assessment"

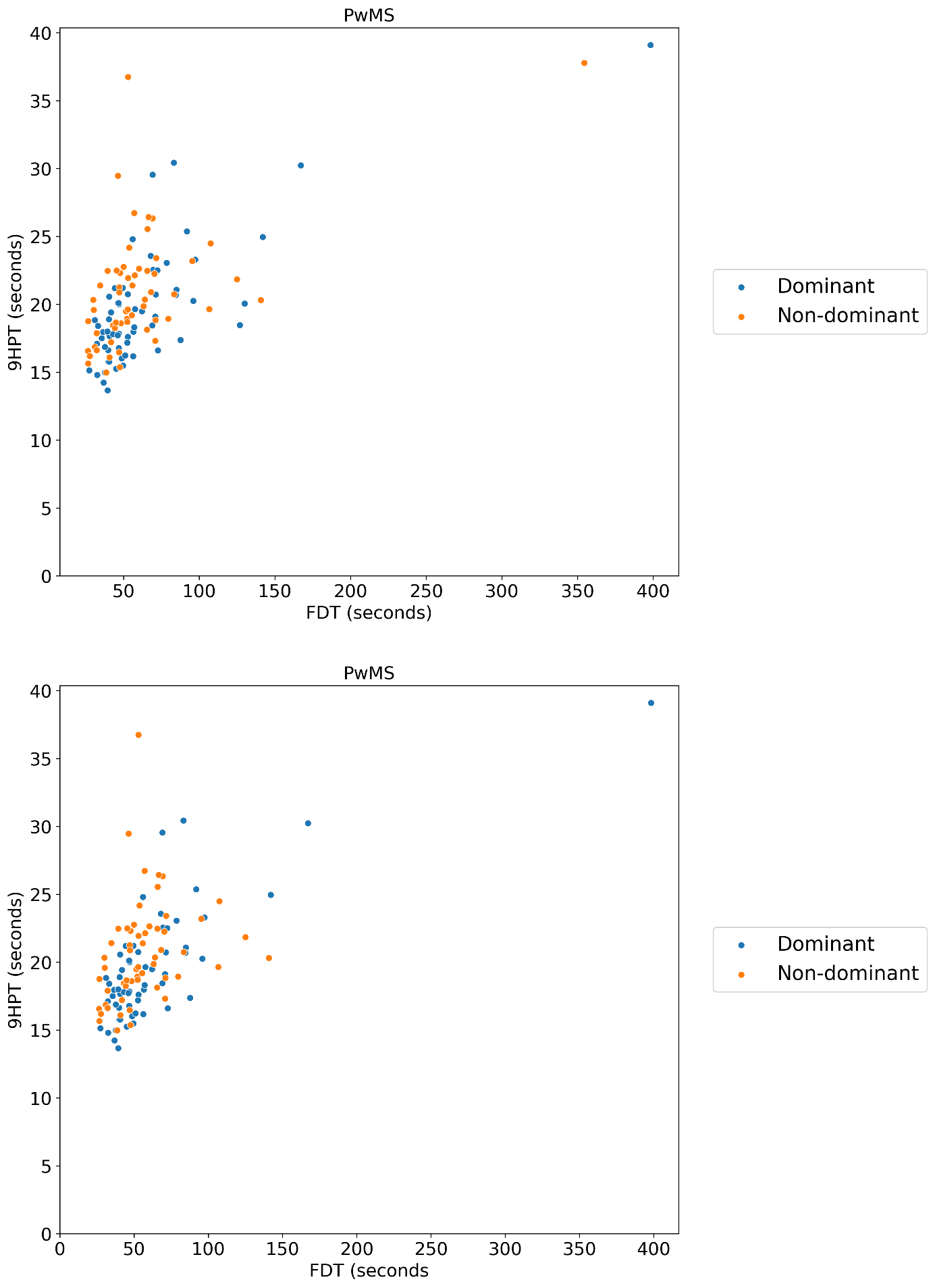

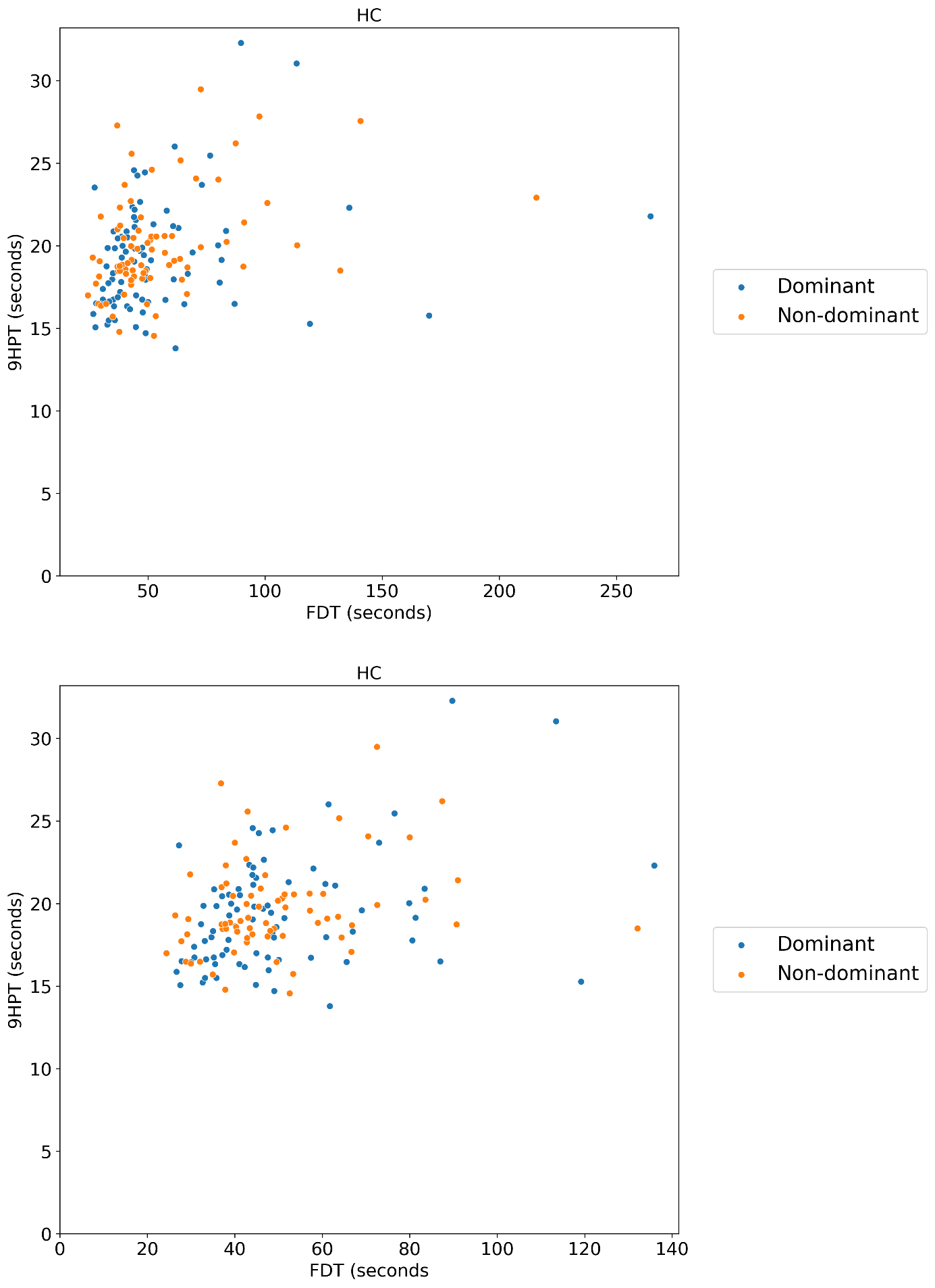

Figure S1 & S2: Concurrent validity - scatterplot between Finger Dexterity Test (FDT) and Nine-Hole Peg Test (9HPT) of subjects with MS (PwMS) and healthy control subjects (HC), without outliers.

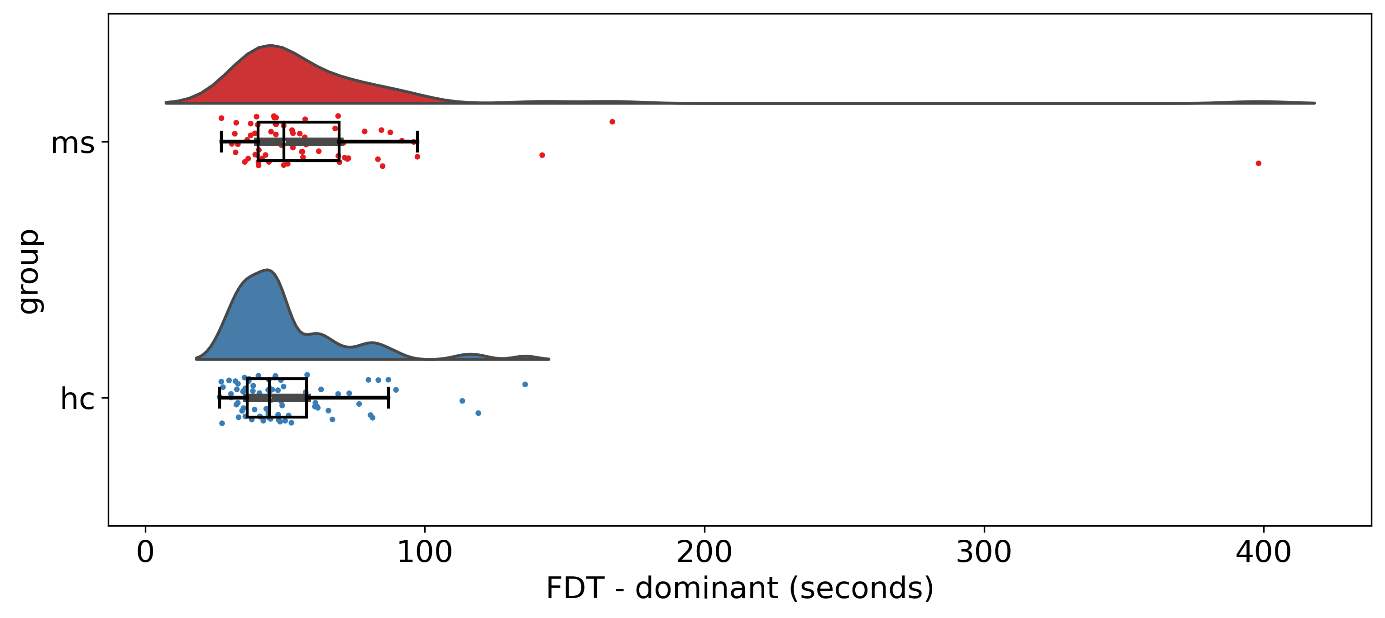

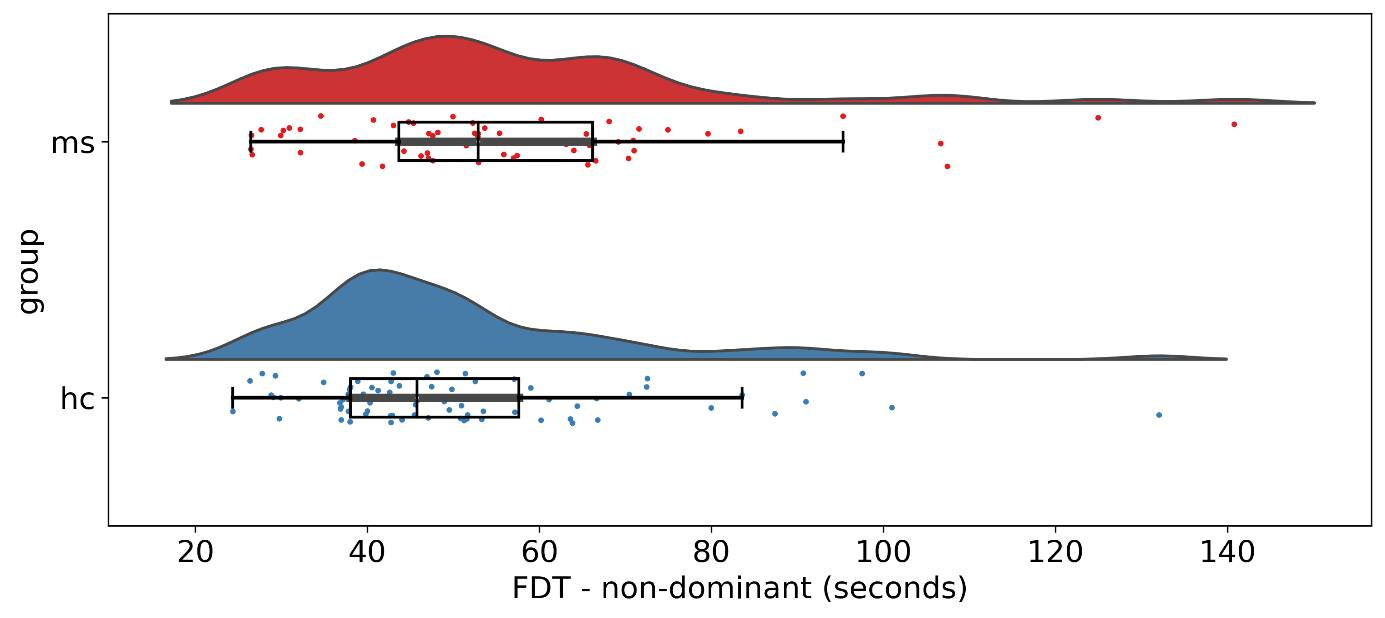

Figure S3 & S4: Group difference of Finger Dexterity Test (FDT) between subjects with MS and healthy controls (HC), for the dominant and non-dominant hand, without outliers

### Regression-based normalisation

To correct performance on the FDT for age and sex (male = 1, female = 2), we performed a regression-based normalisation separately for the dominant and the non-dominant hand. Age and sex were first normalised for both MS and HC data as follows:

$${Variable}_{norm}=\frac{Variable-mean({Variable}_{HC})}{SD({Variable}_{HC})}$$

(eq 1)

We then fitted a linear regression on the HC data:

$$FDT=\beta_{0}+\beta_{1}*Age+\beta_{2}*Sex+\varepsilon$$

(eq 2)

We then used these weights to predict FDT on the HC and MS data and calculate the error between true and predicted FDT:

$${FDT}_{pred}=\beta_{0}+\beta_{1}*Age+\beta_{2}*Sex$$

(eq 3)

$\varepsilon_{pred}={FDT}_{pred}- FDT$

(eq 4)

The final z-score was then calculated as follows:

$$z=\frac{\varepsilon_{pred}}{SD(\varepsilon_{pred,HC})}$$

(eq 5)

The z-score distributions of the dominant and non-dominant FDT can be consulted in figure S5 & S6. After correction for age and sex, the distributions were significantly different between PwMS and HC for the dominant hand (Mann-Whitney U test statistic U = 1741, p = 0.008), but not for the non-dominant hand (U = 1964, p = 0.077).

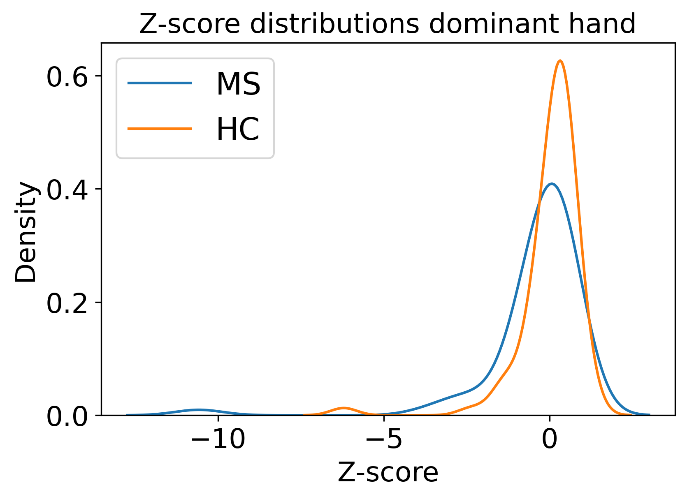

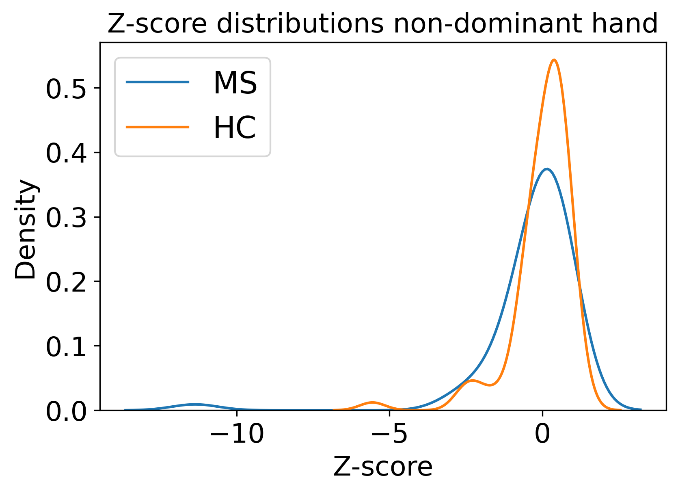

Figure S5 & S6: Z-score distributions for between subjects with MS and healthy control subjects in the dominant and non-dominant hand

The weights to calculate *FDT_pred_* in equation 2 (cfr. supra) can be consulted in table S1.

|  | Dominant | Non-dominant |
| --- | --- | --- |
| N_HC_ | 79 | |
| Mean age_HC_ (SD) | 46.36 (14.83) | |
| Mean sex_HC_ (SD) | 1.65 (0.48) | |
| *β_0_* | 0 | 0 |
| *β_1_* | 0.3559 | 0.4047 |
| *β_2_* | 0.0632 | 0.1558 |
| $SD(\varepsilon_{pred,HC})$ | 0.9408 | 0.9013 |

Table S1: The weights to calculate FDT_pred_ in equation 3 -
SD = standard deviation
